# A longitudinal survey for genome-based identification of SARS-CoV-2 in sewage water in selected lockdown areas of Lahore city, Pakistan; a potential approach for future smart lockdown strategy

**DOI:** 10.1101/2020.07.31.20165126

**Authors:** Tahir Yaqub, Muhammad Nawaz, Muhammad Zubair Shabbir, Muhammad Asad Ali, Imran Altaf, Sohail Raza, Muhammad Abu Bakr Shabbir, Muhammad Adnan Ashraf, Syed Zahid Aziz, Sohail Qadir Cheema, Muhammad Bilal Shah, Saira Rafique, Sohail Hassan, Nageen Sardar, Adnan Mehmood, Muhammad Waqar Aziz, Sehar Fazal, Nadir Khan, Muhammad Tahir Khan, Muhammad Moavia Attique, Ali Asif, Muhammad Anwar, Nabeel Ahmad Awan, Muhammad Usman Younis, Muhammad Ajmal Bhatti, Zarfishan Tahir, Nadia Mukhtar, Huda Sarwar, Maaz Sohail Rana

## Abstract

Severe acute respiratory syndrome coronavirus 2 (SARS-CoV-2) infections has affected more than 15 million people and, as of 22 July 2019, caused deaths of more than 0.6 million individuals globally. With the excretion of SARS-CoV-2 in the stool of symptomatic and asymptomatic COVID-19 patients, its genome detection in the sewage water can be used as a powerful epidemiological tool to predict the number of positive cases in a population. This study was conducted to detect SARS-CoV-2 genome in sewage water during the lockdown. Sewage samples, from 28 pre-selected sites, were collected on alternate days from 13-25 July, 2020 from two selected areas [Johar Town (n = 05) and Township (n = 23)], where smart lockdown were implemented by the government authorities on 9^th^ July, 2020. Genomic RNA was extracted and the SARS-CoV-2 was detected and quantified using commercially available kit through Real-Time PCR. Out of 28, sixteen samples were positive on day one while 19, 17, 23, 17, 05 and 09 samples were positive on day 3, 5, 7, 9, 11, and 13, respectively. Results revealed a decreased positivity rate and SARS CoV-2 genome copies in sewage towards the end of lockdown however few sampling sites did not follow a clear pattern indicating the complexities in sewage water based surveillance i.e time of sampling etc. Hourly sampling from two sites for 24 hours also revealed the impact of sampling time on detection of SARS-CoV-2 genome in sewage. Results of current study insinuate a possible role of sewage-based COVID-19 surveillance in monitoring and execution of smart lockdowns.

## INTRODUCTION

The *Coronaviridae* family clusters consist of several large RNA viruses, which share several structural and functional features. However, members of this family (e.g., MERS-CoV, SARS and now the SARS-CoV-2) recognize different cellular receptors and exploit different entry routes, which affect their species specificity and virulence (Meo et al., 2020). The newly emerged SARS-CoV-2 virus caused pneumonia-like illness in Wuhan, China in December 2019. Affected patients had the symptoms of fever, dry cough, respiratory distress, myalgia, gastrointestinal disturbance, organ failure and loss of smell and taste. The World Health Organization (WHO) named the disease as COVID-19 and declared it pandemic (Gorbalenya et al., 2020). The disease transmission in human to human was very high which spread the disease worldwide within two months. Till July 26, 2020 the number of affected people is 16,207,130 while 648,513 people have died of the disease (Worldometer, 2020).

The positive patient of SARS-CoV-2 may remain asymptomatic or start showing symptoms in 2-11 days of contracting the virus with an average incubation period of 6.4 days (Backer et al., 2020). The viral infection can be diagnosed from nasopharyngeal, throat, alveolar lavage, lacrimal, blood and stool samples. Besides replicating in the respiratory tract, the virus proliferates in the glandular epithelial cells of gastrointestinal tract and the patient starts shedding the virus in stool irrespective of showing the symptoms which makes the sewage based detection to be more beneficial in early infection. From all the samples, nucleic acid based diagnostic is the authentic approach which amplifies the viral nucleic acid by detecting the open reading frame 1ab (*ORF1ab*) gene through real-time RT-PCR (R. Liu et al., 2020).

Owing to an immediate neighbour of an epicentre of the emerging virus SARS-CoV-2 and its subsequent spread across many parts of the world including Pakistan, it is very much essential to conduct studies for necessary intervention to mitigate and augment government authorities for the control of the disease. Raw sewage is one of the significant sources of pathogens that enter the environment, especially viruses that are highly stable in environmental conditions. There are several studies in the subject matter from different countries such as China, USA, Italy, Australia, Germany, and the UK etc. which suggest that raw sewage represent a useful matrix to study viruses excreted by human and animal population (Ahmed et al., 2020; Heijnen and Medema, 2011; Hellmér et al., 2014; W. W. Liu et al., 2020). Indeed, sewage monitoring has previously been employed by the WHO for Poliovirus surveillance during its eradication program (Matsuura et al., 2000).

The SARS-CoV-2 has proven itself to be the most devastating across the world including Pakistan, which led the Government of Pakistan to implement complete or smart lockdown since March 24, 2020. The cessation of business activities along with increased cost of disease prevention and treatment resulted in a huge economic and public health loss. This can be evidenced by a rising number of clinical cases and the ongoing economic constraints worldwide; however, the consequences of these constraints are very much significant for developing nations such as Pakistan (Waris et al., 2020). For a resource-limited setting such as Pakistan where a large number of populations relies exclusively on daily wages, masses can’t survive complete lockdown for a longer period. Furthermore, there are concerns of sub-optimal control of the disease in the ongoing lockdown approach based on the wrong data of address and impersonate entries provided by patients due to stigma associated with disease(Murakami et al., 2020).

With this background, in collaboration with the Water and Sanitation Agency (WASA) directorate in the Lahore region, we conducted this particular study to predict sewage-based disease burden in a particular area or setting to facilitate government authorities to intervene either for implementation of an effective lockdown or otherwise. Sewage samples across different sites in two lockdown areas were collected and processed for genome-based detection of SRAS-CoV-2. The gained outcomes and experience is worth replicative to any other setting comparable to the current one worldwide.

## MATERIALS AND METHODS

### Sampling sites and strategy for sewage water

Lahore is the Capital of the province Punjab with an estimated populace of 12,642,423 (World Population, 2020). During the pandemic of SARS-CoV-2, Pakistan Government used smart lock down strategy to combat spread of COVID-19. On 9^th^, July 2020, a two week smart lockdown was implemented in seven areas of Lahore, Pakistan. We selected two areas with smart lockdown (Johar town C Block and Township A II sector) for longitudinal detection and quantification of SARS CoV-2 genome in sewage water of these areas during the lockdown. Sewage water samples were collected from 5 pre-selected sites (01 lift station and 4 sewage lines) of Johar town C Block and 23 sites (19 sewage lines and 04 lift/disposal stations) of Township sector AII using grab sampling method. Sewage water samples were collected on alternate days from lockdown areas from July 13 to 25, 2020. Furthermore, a 24 hours sewage water sampling was done from Bheer Pind lift station, Johar Town and A II lift station Township to determine the effect of sampling time on the detection and quantification of SARS CoV-2 genome. At the time of sewage sampling, sampling personnel used standard personal protective equipment, such as long pants, boots, hats, safety glasses and gloves. Samples were transported to the BSL-3 facility at the Institute of Microbiology (IM), University of Veterinary and Animal Sciences (UVAS) Lahore, Pakistan in a cool box and stored at 4 °C until further analysis within 24 hours.

### RNA Extraction from Sewage Samples

RNA of each sewage sample was extracted in BSL-3 of IM, UVAS Lahore, Pakistan. Before extraction, each sample was vortexed thoroughly and 1 ml of the sample was transferred to microfuge tube. Samples were centrifuged at 5000 rpm for 15 minutes at 4 °C. The supernatant was used for RNA extraction. RNA was extracted using the Hero 32 extraction system. A 14 µL of Proteinase K + Carrier RNA mixture was added into each well of RNA extraction plate. Then 200 µL of each of the supernatants was added into the respective wells. The extraction plates were placed in RNA extractor and the program was run. RNA was extracted from the elution wells of plates and stored at −80°C and subjected to RT-qPCR analysis at the same day. In order to reduce the potential RT-qPCR contamination, RNA extraction and RT-qPCR setup were performed in separate laboratories.

### RT-qPCR analysis

RT-qPCR analysis of sewage samples was performed by using the commercially available kit (2019-nCoV Nucleic Acid Diagnostic Kit, Sansure Biotech Inc., China). We used Open Reading Frame 1ab (ORF1ab) gene and nucleocapsid protein (N) gene for SARS-CoV-2 detection by RT-qPCR. Each reaction mixture contained 2019-nCoV-PCR Mix (26µl), 2019-nCoV-PCR Enzyme Mix (4µl) and 20 µl RNA template to make the final volume of 50 µl. Thermal cycling reactions were carried out at 50 °C for 30 minutes, followed by 95 °C for 1 min and 45 cycles of 95 °C for 15 and 60 °C for 30 seconds on CFX-96 Real Time PCR detection system (Bio-Rad Laboratories). All RT-qPCR reactions also had positive and negative controls. Reactions were assumed positive if the cycle threshold was below 40 cycles. To calculate the SARS CoV-2 genome copies in sewage, a standard curve based on ORF1ab gene was generated from different dilutions of positive control of kit (Figure 1).

**Figure 1:**
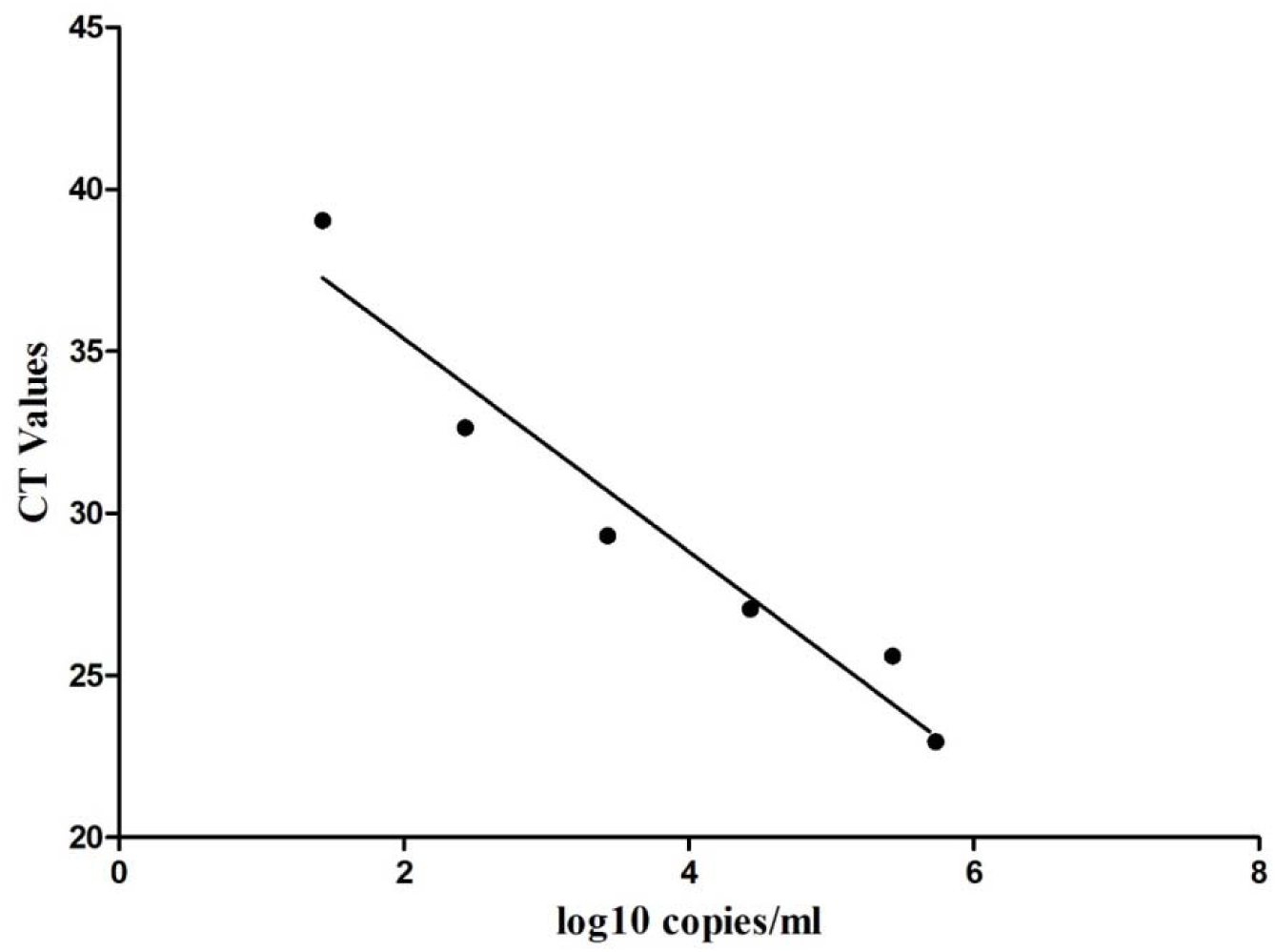
Standard curve of CT values of real-time RT-PCR against the log 10 copies/ml of SARS-CoV-2.

## RESULTS

A total of 196 sewage samples were collected from 28 sites (07 samples from each site) including 05 sewage sites of C-2 Block Johar town and 23 sewage sites of C-2 Block Township, Lahore, Pakistan on alternate days from 13^th^ July, 2020 to 25^th^ July, 2020. The longitude and latitude of sampling sites of C-Block, Johar Town and A-II sector, Township area are displayed in the Table 1. Out of 28, sixteen samples were positive on day one while 19, 17, 23, 17, 05 and 09 samples were positive on day 3, 5, 7, 9, 11, and 13, respectively (Table 2, Figure 3). Out of 05 sampling sites of C-Block, Johar Town, 04 sites were street sewerage lines and 5^th^ site was relevant sewage lift station. All street sewage pipelines of Johar Town had 9 inches diameter. On the first day of sampling (13^th^ July), all street sewage pipe lines (4/4) were detected positive for the SARS-CoV-2 with variable load of SARS CoV-2 genome (10^2.426 to 10^4.556) (Table 01; Figure 4). Sampling site 01 remained positive for SARS CoV-2 genome throughout the study with highest genome copies 10^3.64 on 4^th^ sampling on 19^th^ July, 2020. The SARS CoV-2 genome was not detected from subsequent samples from sites 2-4 throughout the study period except 4^th^ and 5^th^ sample from site 03 (Table 1). The SARS CoV-2 was not detected from the lift station of Johar Town (site-05) on first sampling on 13^th^ July while two subsequent samples on 15 and 17 were detected positive for SARS CoV-2 with a decreasing viral load (10^2.73 and 0.60 copies/ ml, respectively). Overall, towards the end of lock down on 23^rd^ July, SARS CoV-2 was not detected from 80% (4/5) sewage sites of C-Block Johar town.

**Table 1:** SARS-CoV-2 RNA copies in sewage water samples of two different areas of Lahore under lockdown.

| Johar Town | Line diameter (inches) | Longitude | Latitude | 1 | 2 | 3 | 4 | 5 | 6 | 7 |
| --- | --- | --- | --- | --- | --- | --- | --- | --- | --- | --- |
|  |  |  |  | log10 SARS CoV-2 RNA copies/ml of sewage |  |  |  |  |  |  |
|  |  |  |  | 13/07 | 15/07 | 17/07 | 19/7 | 21/7 | 23/7 | 25/7 |
| Site 01 | 9 | 74.29545 | 31.4524723 | 3.03 | 2.43 | 2.730 | 3.643 | 1.82 | 1.51 | 2.73 |
| Site 02 | 9 | 74.2977366 | 31.4557036 | 3.94 | 0 | 0 | 0 | 0 | 0 | 0 |
| Site 03 | 9 | 74.2942093 | 31.4573861 | 2.42 | 0 | 0 | 0.91 | 1.82 | 0 | 0 |
| Site 04 | 9 | 74.2989921 | 31.4552673 | 4.55 | 0 | 0 | 0 | 0 | 0 | 0 |
| Bheer Pind L/S | 42 | 74.2952135 | 31.4572428 | 0 | 2.73 | 0.60 | 0 | 0 | 0 | 0 |
| **Township AII Sector** |  |  |  |  |  |  |  |  |  |  |
| Site 01 | 9 | 74.2928192 | 31.435475 | 0.60 | 0.91 | 0.60 | 1.11 | 0.91 | 2.12 | 0 |
| Site 02 | 12 | 74.294026 | 31.454058 | 3.64 | 3.03 | 1.21 | 0 | 1.82 | 0 | 0 |
| Site 03 | 36 | 74.2929095 | 31.4352528 | 3.95 | 0 | 0 | 1.24 | 0 | 0 | 0 |
| Site 04 | 36 | 74.2929237 | 31.4354824 | 0 | 2.43 | 0 | 0 | 0 | 0 | 0.91 |
| Site 05 | 9 | 74.3165282 | 31.4551185 | 2.73 | 0 | 0.91 | 0.72 | 0 | 0 | 0 |
| Site 06 | 9 | 74.3157992 | 31.4552004 | 3.34 | 2.73 | 0.91 | 2.85 | 0 | 0.60 | 0 |
| Site 07 | 9 | 74.3175337 | 31.4546141 | 0 | 2.73 | 0 | 2.48 | 0.60 | 0 | 0 |
| Site 08 | 18 | 74.3180418 | 31.4547766 | 0 | 2.43 | 1.21 | 3.40 | 0.30 | 0 | 0 |
| Site 09 | 18 | 74.3166218 | 31.4552856 | 0 | 0.91 | 0 | 0.97 | 0.91 | 1.21 | 0 |
| Site 10 | 18 | 74.3175544 | 31.4549163 | 0 | 0.91 | 0 | 1.149 | 1.51 | 0.90 | 0 |
| Site 11 | 9 | 74.3175511 | 31.4546758 | 2.43 | 0.91 | 0 | 4.00 | 0.91 | 0 | 0 |
| Site 12 | 9 | 74.3189623 | 31.451292 | 0.91 | 0.91 | 3.03 | 1.45 | 0 | 0 | 0 |
| Site 13 | 9 | 743191111 | 31.4515717 | 0 | 0.91 | 0 | 3.58 | 0 | 0 | 0 |
| Site 14 | 9 | 74.3145578 | 31.4526552 | 0 | 0.91 | 0 | 0 | 0 | 0 | 0 |
| Site 15 | 9 | 74.3142842 | 31.4521259 | 0.91 | 3.95 | 1.21 | 0.97 | 0.60 | 0 | 2.43 |
| Site 16 | 9 | 74.3157992 | 31.4552004 | 0 | 0.60 | 0.60 | 0.84 | 2.12 | 0 | 0.60 |
| Site 17 | 9 | 74.3165282 | 31.4551185 | 0.60 | 0.60 | 0.60 | 3.34 | 0 | 0 | 0.91 |
| Site 18 | 9 | 74.3145578 | 31.4526552 | 3.34 | 0.78 | 0.60 | 1.00 | 2.43 | 0 | 3.64 |
| Site 19 | 9 | 74.3175337 | 31.4546141 | 0 | 0 | 0.60 | 0.72 | 2.73 | 0 | 0 |
| Township A II LS | 36 | 74.3128196 | 31.4515134 | 0 | 3.34 | 0.60 | 2.43 | 0.91 | 0 | 0.91 |
| C2, Green Town DS | 36 | 74.3125476 | 31.455127 | 1.82 | 0 | 0.90 | 3.70 | 1.51 | 0 | 3.03 |
| Ameer Chowk, DS | 36 | 74.289159 | 31.4343771 | 0 | 0 | 3.31 | 2.43 | 1.51 | 0 | 0 |
| Sadiq Chowk, C1, DS | 36 | 74.3055126 | 31.4483829 | 1.21 | 0 | 0.60 | 1.51 | 1.51 | 0 | 0.60 |

LS: lift station; DS: Disposal station

**Figure 2.**
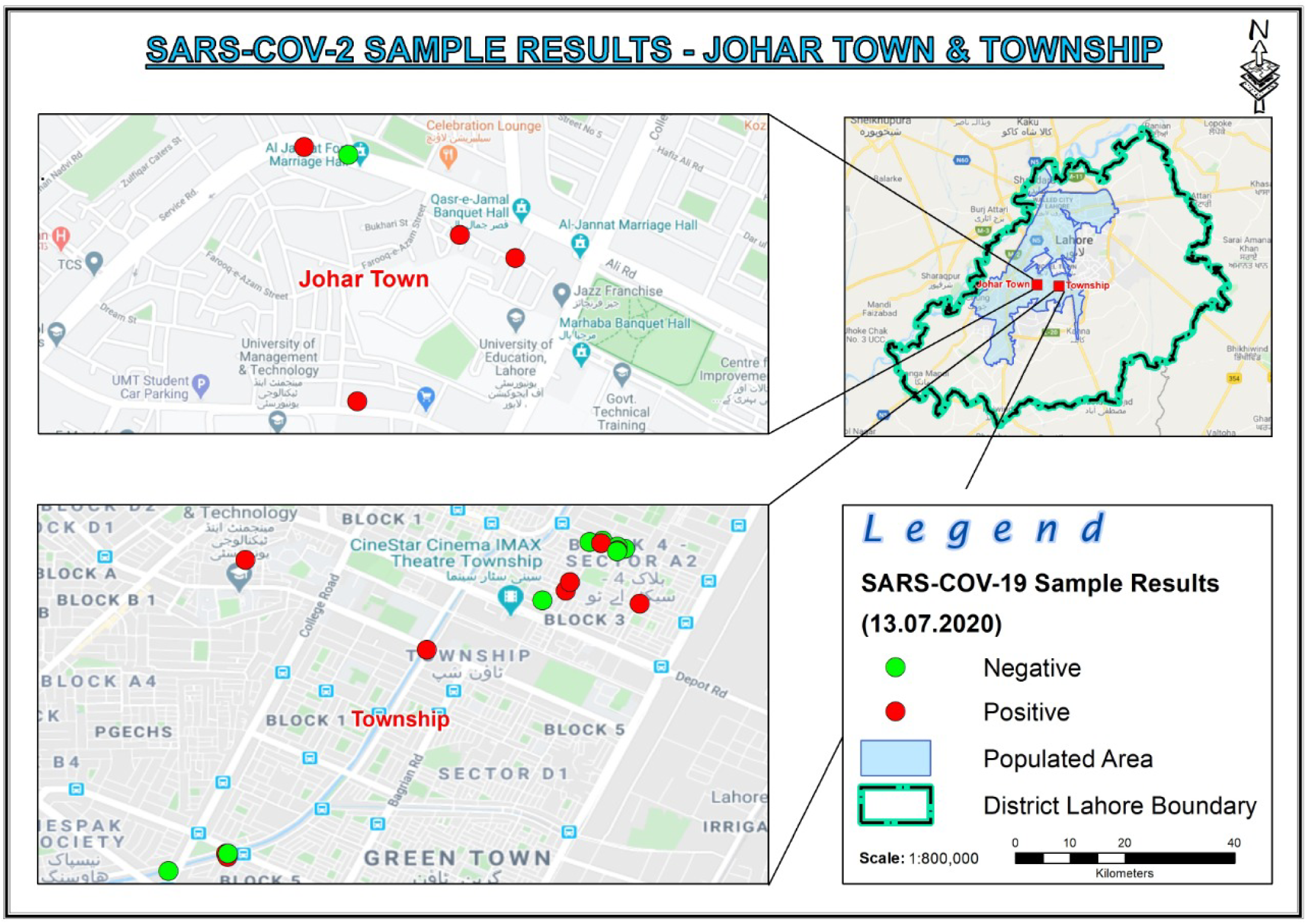
Satellite map of Johar Town and Township areas. Red dots indicate the SARS-CoV-2 positive samples, while Green dots are the SARS-CoV-2 negative samples of smart lockdown areas on 13^th^ July, 2020.

**Figure 3.**
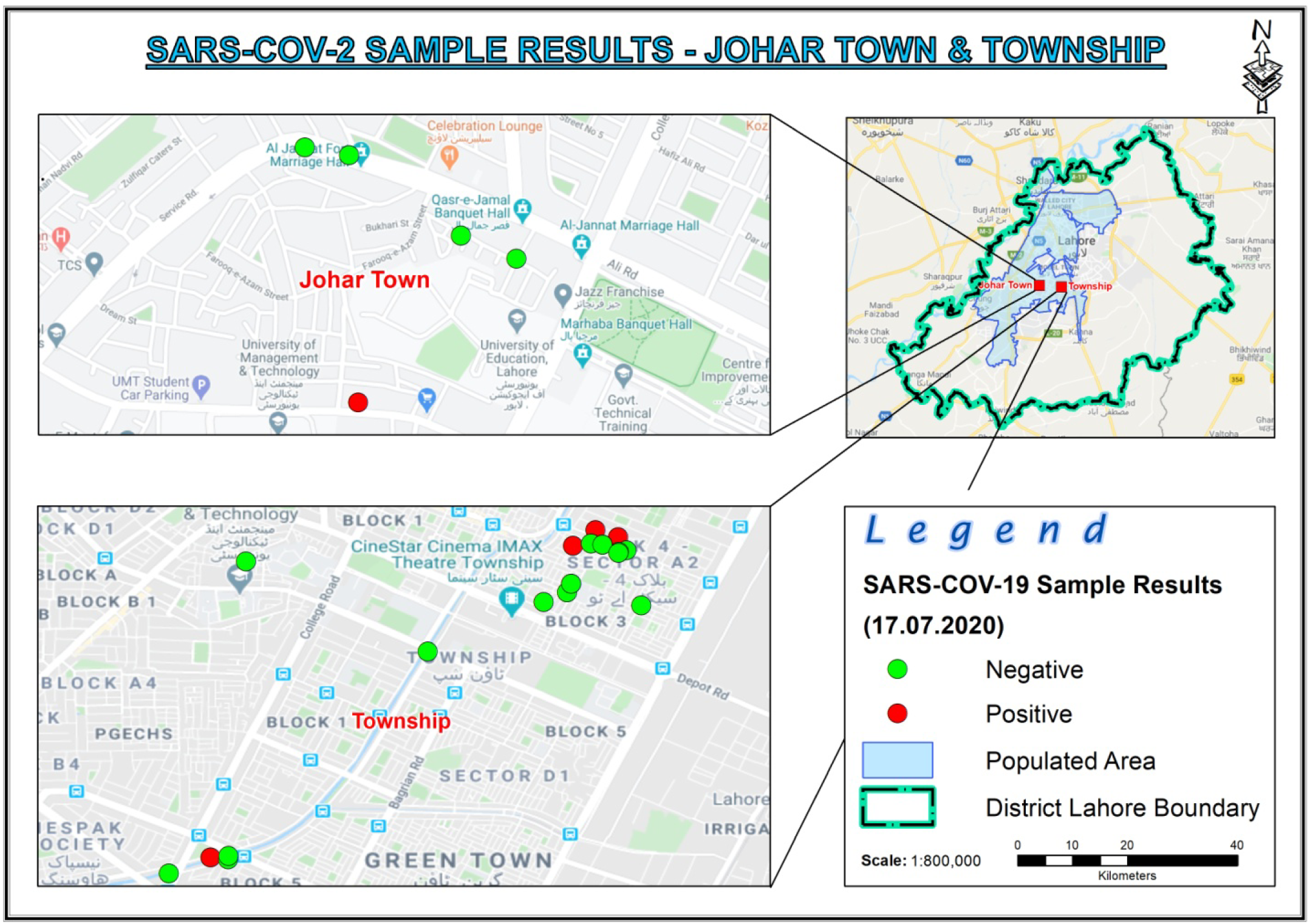
Satellite map of Johar Town and Township areas. Red dots indicate the SARS-CoV-2 positive samples, while Green dots are the SARS-CoV-2 negative samples of smart lockdown areas on 23^rd^ July, 2020.

**Figure 4:**
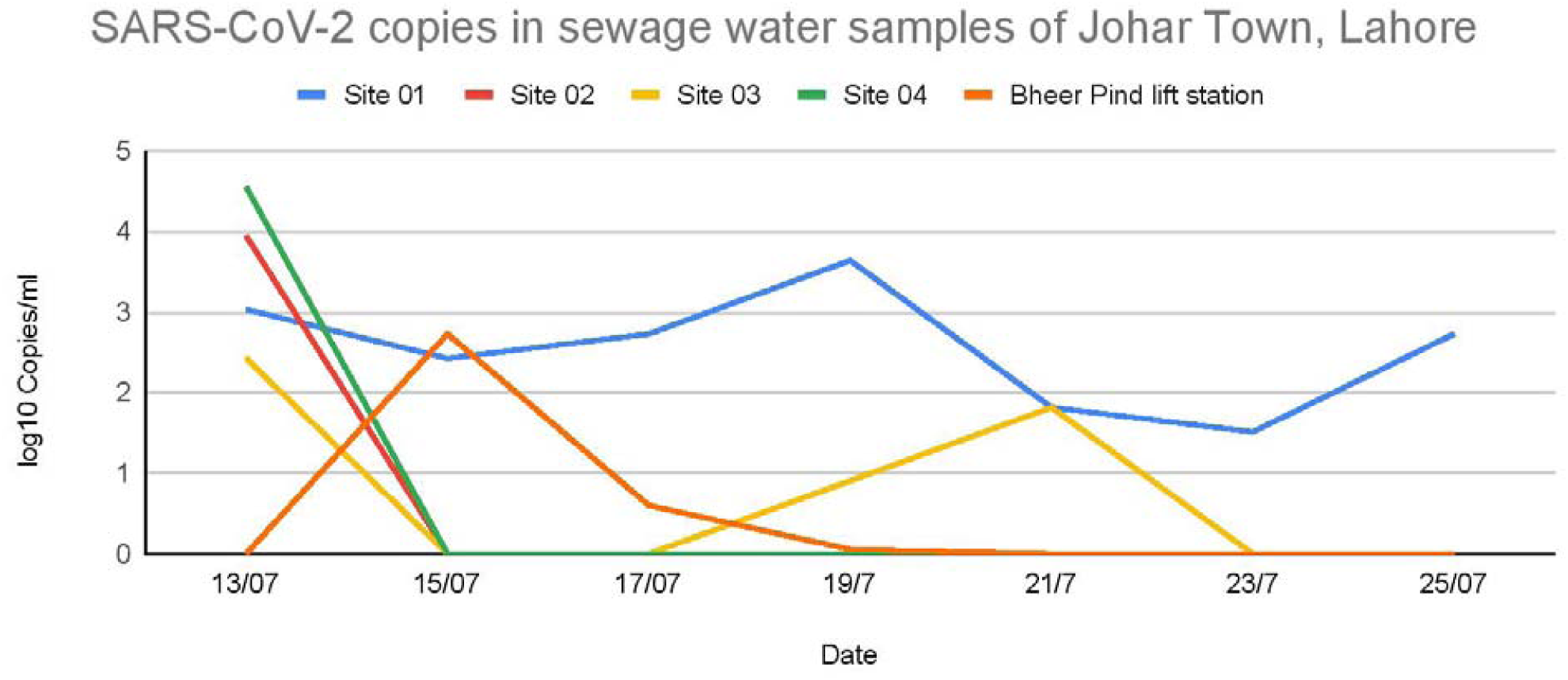
SARS-CoV-2 genome load as log 10 copies/ ml at the sites of C Block, Johar town with respect to progression of lockdown days.

A total 23 sewage sites were sampled from the A-2 Block of Township, Lahore based on the population density and sewage maps of area. Among these sites, 19 sites were street sewage pipelines of different diameters i.e 9, 12, 18 inches while remaining 04 sites were relevant downstream lift/disposal stations. Out of 19 sites of street sewage pipelines, 10 were detected positive for SARS CoV-2 on first sampling on 13^th^ July while 16/19, 11/19,16/19,11/19, 04/19 and 05/19 sites were detected on subsequent samplings on 15^th^,17^th^, 19^th^, 21^st^, 23^rd^ and 25^th^ July,2020, respectively (Table 01, Figure 5). Among 4 lift/disposal stations of Township area, 2/4 were detected positive on first sampling while all four stations were detected positive on 3^rd^, 4^th^, and 5^th^ sampling with higher load of SARS CoV-2 genome. Toward the end of lockdown all four lift/disposal stations were detected negative for SARS CoV-2. To access the role of sampling time on detection of SARS CoV-2 in sewage, two sewage lift stations, one from each area under smart lock down, were sampled on hourly basis for 24 hours and detection of SARS CoV-2 from hourly samples is presented in Figure 6. Different sampling time was revealed to be better for both areas.

**Figure 5:**
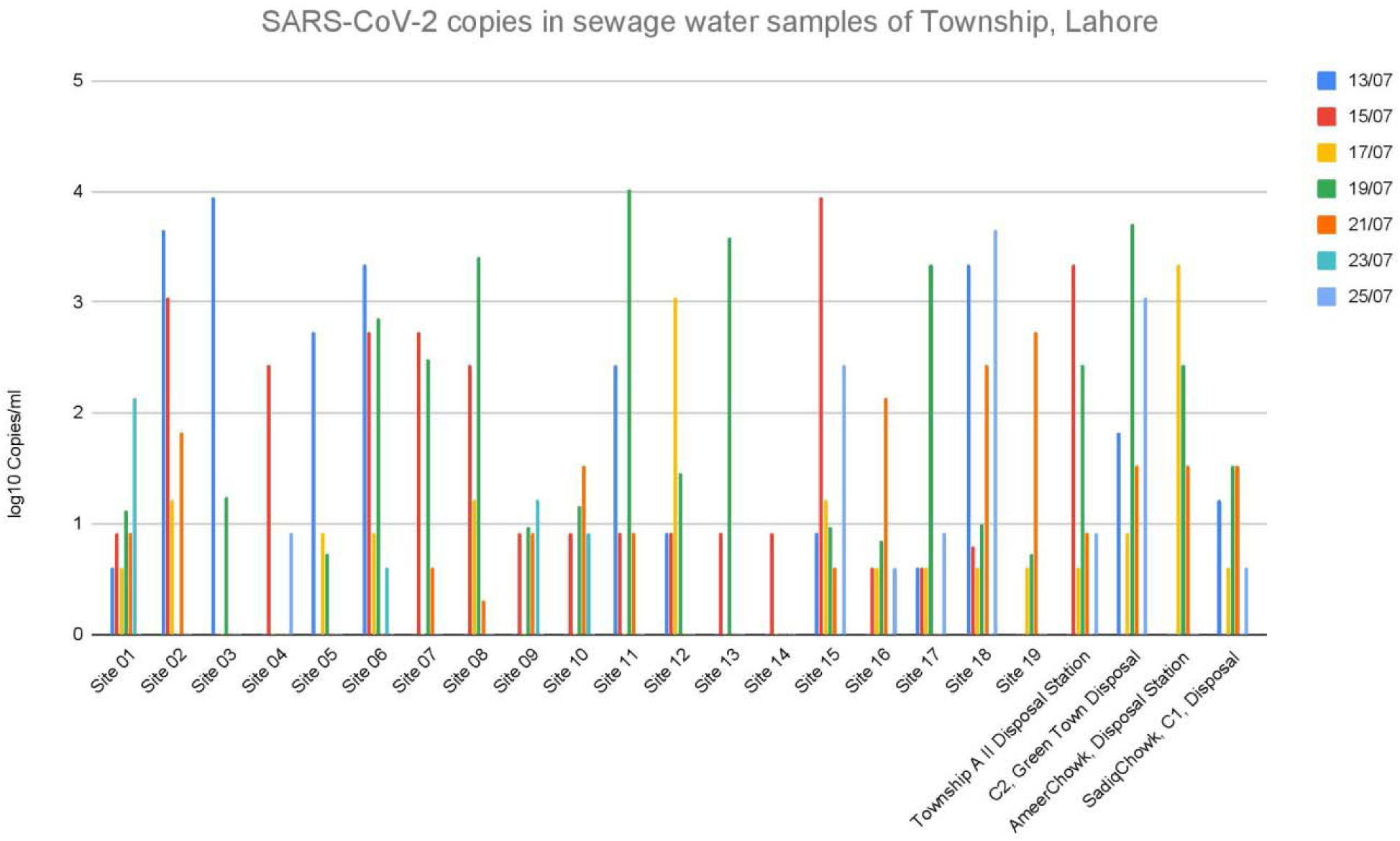
Graphical display of the SARS-CoV-2 viral load as log 10 copies/ ml at the sites of Township with respect to progression of lockdown days.

**Figure 6:**
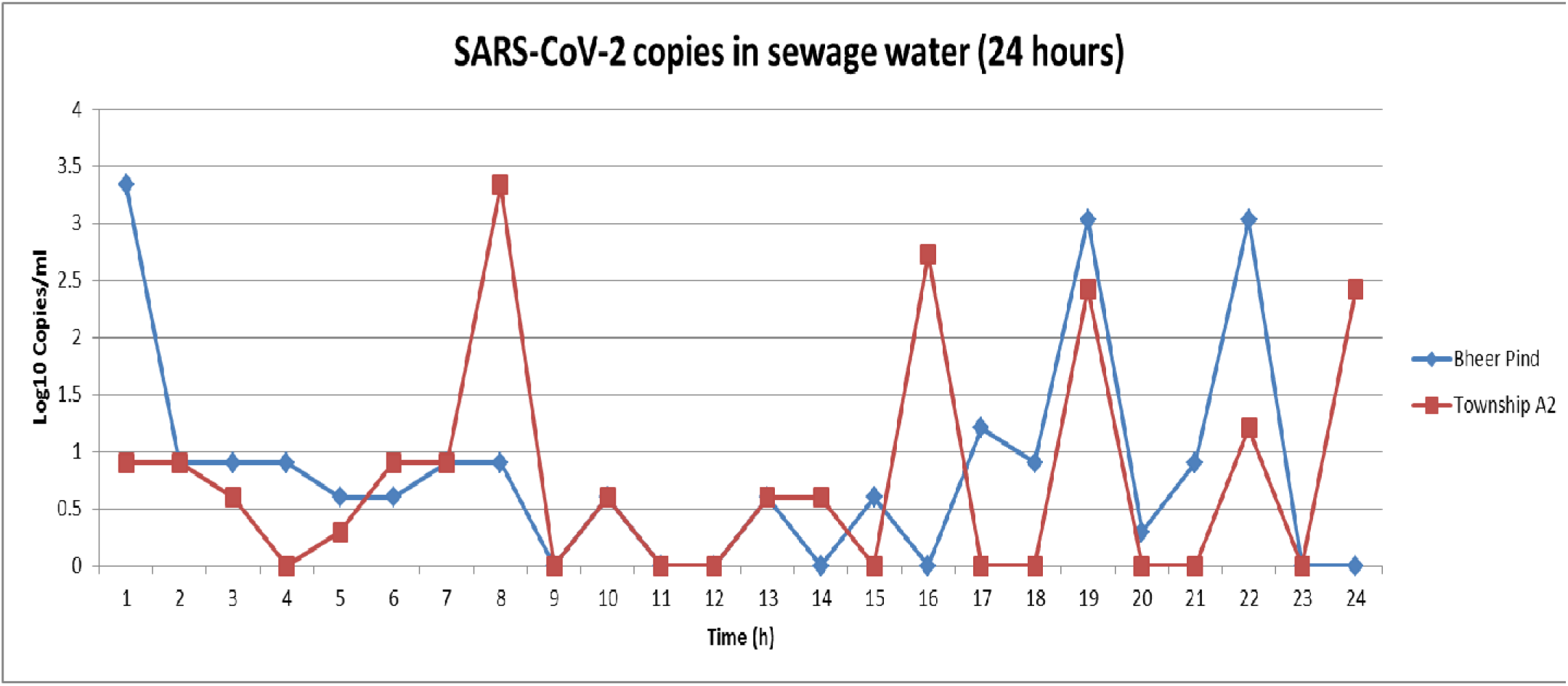
Graphical display of the SARS-CoV-2 viral load as log 10 copies/ ml at Site 05 of Johar Town and Site 20 of Township with respect to time in hours starting from 1:00 am for the period of 24 hours.

## DISCUSSION

The short incubation period and the shedding of virus from the infected as well as the asymptomatic cases allowed the rapid spread of the virus. Medical screening of the symptomatic patients is a biased method to estimate the prevalence of the disease because asymptomatic cases are the major portion of the COVID-19 patients. Most of the developing countries remain under-tested due to resources thus disease occurrence remains underreported. However, some developed countries are testing huge number of individuals to obtain population-wide data such as china, USA and Russia but, this approach is slow and requires more resources (Hart & Halden, 2020). Therefore, a more practical approach should be tested that not only be economical but also practical. Waste water-based epidemiology (WBE) has been identified previously a good tool for disease surveillance with a good proven track record in hepatitis A and polio (Asghar et al., 2014; Hellmer et al., 2014). SARS-CoV-2 has been detected in fecal samples of both symptomatic and asymptomatic patents of CoVID-19 patients, previously (Cai et al., 2020; Gao, Chen, & Fang, 2020; Holshue et al., 2020; Tang et al., 2020; Wolfel et al., 2020). Epidemiologists believe WBE may prove a good strategy for population based-surveillance of the COVID-19. Real time PCR is considered to be an optimum test for identification of SARS-CoV-2 (Carter et al., 2020). Moreover, reduced biosafety requirements and the cost have made RT-PCR a good choice to be used for diagnosis and WBE of SARS-CoV-2. Current reports of successful SARS-CoV-2 detection in human stool samples and the waste water proves that WBE may prove to be a good tool for disease surveillance in the population. The WBE of COVID-19 will also help in the strategies of imposing or lifting off lock down/smart lock down in an area.

A longitudinal study was planned to detect SARS-CoV-2 RNA in the untreated wastewater samples of two lock down areas of the Lahore, Pakistan. Lahore is a densely populated city with a population size of more than 12 millions. The dense population areas have high COVID-19 infection rate and death rate (Shima Hamidi 2020). These lock-down areas represent the typical urban settings where people of different classes and from different backgrounds live. The wastewater samples were collected from the sewerage lines on alternate for fourteen days. In this study, we used commercially available Sansure kit for viral genome detection. The detection efficacy of SARS-CoV-2 genome by Sansure kit is higher than BGI and Bioperfectus (Lihua Shen 2020). Previous studies used different viral concentration methods to concentrate the viral RNA from the waste water (Masaaki Kitajimaa 2020). In current study, we report SARS CoV-2 detection from sewage of lockdown areas with concentrating the sample. First sampling from pre-selected sites was done on 4^th^ day of implementation of lock down and SARS CoV-2 RNA was detected from majority of sampling sites from both areas. Towards the end of lockdown on 23^rd^ July, only a small number (5/28) of sampling sites were detected positive. Some of the sampling sites did not showed a clear pattern which may be attributed to the time of sampling and changing disease pattern within area or movement of asymptomatic carriers within area. Our results also indicate that sampling time is very crucial for sewage based surveillance of COVID-19 in an area and best time of sampling may vary between different localities depending on different factors. The results showed that in both areas viral genome was detected in early morning (7 AM to 10 AM) and evening time to mid night (6PM to 12 PM). This may be due to peaks in toilet use in the morning and evening (Coutu et al., 2013). In Pakistan, these are the times when most of the working people are at home. These results suggest that hour to hour variation in viral genome detection in waste water of same sampling site affects the final conclusion therefore; before sampling appropriate sampling time should be selected by using hourly sampling method. Detection of SARS-CoV-2 in waste water samples for a period of time clearly explain that sewage-based surveillance could be employed for implementation and lifting of smart lock-down and routine surveillance. Sewage-based surveillance system can predict the number of symptomatic and asymptomatic patients in a locality. This study is still ongoing and we are in process of further data analysis and establishing a link between reported COVID 19 cases in these areas and detection of SARS CoV-2 from sewage samples of these areas. Sites which were persistently positive for SARS CoV-2 genome are still under surveillance. One of the striking observations of study is that SARS-CoV-2 RNA can be detected in raw sewage water samples without concentrating the virus, at least in lockdown areas where a higher disease burden is expected. The affect of sewage water flow rate and environmental factors like rain on SARS-CoV-2 RNA detection is a big challenge in the sewage – water based surveillance. Collectively, this study provides an indication of sewage-water based COVID-19 surveillance to predict the disease prevalence and burden in an area and a possible role in monitoring and execution of smart lockdowns.

## Data Availability

All data is available in the article. Raw file can be provided on request

## ACKNOWLEDGMENTS

We thank Government of Punjab for financial support to this project and the WASA directorate Lahore region in facilitation of sampling across the district. We also thank laboratory technicians and support staff in the Institute of Microbiology throughout the sampling and sample processing work in the BSL-3 facility.

